# Association of Circulating Calcitonin With Risk and Onset of Postoperative Atrial Fibrillation After Cardiac Surgery

**DOI:** 10.64898/2026.05.14.26353191

**Authors:** Chi Him Kendrick Yiu, Lucia M. Moreira, Ioannis Akoumianakis, Peter Rothwell, Charalambos Antoniades, Svetlana Reilly

## Abstract

**Background:** Postoperative atrial fibrillation (POAF) affects up to 50% of cardiac surgery patients and is linked to higher morbidity, longer hospital stays and increased thromboembolic risk. Early identification of at-risk patients remains challenging. Calcitonin (CT), a hormone with anti-fibrotic effects, may serve as a novel biomarker.

**Methods:** In 491 patients undergoing elective cardiac surgery, baseline serum CT was measured preoperatively using CT-specific enzyme-linked immunosorbent assay (ELISA). Patients with pre-existing AF were excluded. Associations between CT levels and POAF incidence and onset were evaluated using logistic regression, Cox proportional hazards models, and Kaplan–Meier analysis.

**Results:** Among 248 patients with detectable CT levels, 88 patients developed POAF. Higher baseline CT was independently associated with lower risk of POAF (OR 0.68 per 5 pg/ml increase; 95% CI 0.51-0.89; *P* = 0.009) and delayed arrhythmia onset (adjusted HR 0.941; 95% CI 0.898-0.980, *P* = 0.0026) after adjusting for covariates. Kaplan–Meier analysis demonstrated a graded relationship between increasing CT levels and reduced cumulative incidence of POAF. In this cohort, baseline CT showed greater discriminative ability than CRP and BNP, although overall model performance remained moderate.

**Conclusion:** Higher preoperative circulating CT levels are associated with reduced risk and delayed onset of POAF following cardiac surgery. These findings suggest that calcitonin may have the potential as a biomarker for perioperative risk stratification in POAF. Given the observational design and single-centre setting, further validation in independent cohorts and studies integrating mechanistic insights are warranted.

## Introduction

Postoperative atrial fibrillation (POAF) is one of the most common complications following major surgery, affecting 20%-55% of patients depending on the procedures, with higher rates (40-50%) observed after valvular surgery [1, 2]. POAF typically occurs within the first 2-4 days after surgery, however the arrhythmia can develop at any point during the early recovery period [3].

Although often considered a transient and self-limiting arrhythmia, POAF is associated with significant clinical outcomes, including prolonged hospitalization, acute heart failure, myocardial infarction, stroke, cardiac arrest and increased long-term mortality [1, 4]. Despite its high prevalence and clinical impact, the pathophysiology of POAF remains incompletely understood. POAF is recognised as a multifactorial condition arising from the interaction between pre-existing atrial substrate and acute postoperative triggers [2], advanced age, valvular surgery atrial enlargement and pre-existing heart failure [1, 2]. Moreover, inflammatory activation plays a central role, driven by surgical trauma, cardiopulmonary bypass, and ischemia-reperfusion injury, leading to transient atrial electrical instability [3]. However, current clinical risk prediction models incorporating these factors demonstrate only modest discriminatory ability [5, 6]. This limited predictive accuracy highlights the need for novel biomarkers that can capture the underlying pathophysiological processes and identify high risk patients before arrhythmia onset.

Calcitonin (CT) is a peptide hormone classically produced by the thyroid gland to regulate calcium homeostasis [7], but it is also produced locally in atrial myocardium, where it exerts cardioprotective and anti-fibrotic effects [8]. Specifically, CT exerts anti-fibrotic and anti-arrhythmic effects by suppressing collagen deposition and fibroblast proliferation in the atria [8]. While the local cardioprotective roles of cardiac CT have been characterized in experimental studies, the contribution of circulating CT to AF risk, particularly in the early postoperative setting, remains unexplored. In particular, the potential contribution of serum CT to early-stage AF pathogenesis, including POAF, remains unclear. Understanding whether baseline circulating CT levels are associated with the development of POAF may provide valuable insights into disease mechanisms and reveal a novel biomarker for risk prediction.

Given the substantial clinical and economic burden of POAF, the limitations of current predictive models, and the emerging evidence for CT’s cardioprotective properties, there exists a compelling rationale to investigate whether circulating CT levels may serve as a novel biomarker for POAF risk stratification. Therefore, we investigated whether preoperative serum CT levels are associated with (i) the risk of developing POAF and (ii) the timing of arrhythmia onset in patients undergoing elective cardiac surgery. The overall goal of the study was to identify new avenues for POAF early detection and preventive interventions.

## Methods

### Study population and design

491 patients were enrolled in the AdipoRedOx sub-study of the Oxford Cohort for Heart, Vessels and Fat (OxHVF). The study was approved by the local ethics committee (REC 11/SC/0140) at John Radcliffe Hospital, Oxford, and all protocol procedures were in agreement with the declaration of Helsinki. Patients were eligible for this study if they fulfilled the following inclusion criteria: adults, scheduled for elective cardiac surgery and able to provide written informed consent. Exclusion criteria include patients with active infection or on active cancer treatment. In this study, patients with a medical history of prevalent AF were excluded for analysis. POAF was defined as the patients who develop episodes of AF during the postoperative hospitalisation period (information retrieved from the medical notes with confirmation of the diagnosis on at least one 12-lead ECG). The day of POAF onset or diagnosis was documented for each patient if available.

### Blood collection and processing

Whole fasting blood was routinely collected from patients on the morning of surgery. Subsequently, blood was transferred to BD Vacutainer Yellow SST II advance tubes. Samples were left for 1 hour at room temperature to clot, followed by centrifugation at 3000 rpm for 15 minutes at room temperature. After separation of the layers, serum was collected from the top layer and kept at -80℃ for long term storage until use.

### Data collection and outcomes

Clinical data were collected from the electronic patient record at baseline and followed up until patients were discharged from hospital. Data included demographic characteristics, surgical procedure, medical history, medications and postoperative events. The following information were also recorded if available: biochemical measurements, echocardiography measurements. The primary outcomes of this study were the postoperative rhythm status (POAF or sinus rhythm) and the day of onset in patients with POAF. Secondary outcomes were clinical events after surgery, including inotropes duration, bleeding in hospital, fresh frozen plasma (FFP) requirement, platelet units given, red blood cell (RBC) requirement, mechanical ventilation hours, days in hospital and postoperative death.

### CT measurement

CT levels in the serum samples were measured using the high-sensitivity enzyme-linked immunosorbent assay (ELISA) (#RIS0019R, BioVendor) according to manufacturer’s protocol. In brief, 100 μl of each calibrator, control or sample were loaded to the wells of the CT-coated microtiterplate. A working anti-CT-HRP conjugate solution (50 μl) was then added to all wells and incubated for 18 hours at 4℃, followed by three washing steps with a wash solution (400 μl). Next, 100 μl of chromogenic solution was added and incubated for 30 minutes at room temperature in the dark. Finally, 100 μl of stop solution were added and absorbance was measured at 450 nm using the FLUOstar Omega microplate reader. The concentration of CT was expressed in pg/ml. Specificity of the CT ELISA kit was validated in **Figure S1**.

### Statistical analysis

Statistical analysis was performed using GraphPad Prism 10.5.0 and R version 4.2.3. Baseline characteristics were compared between the sinus rhythm and POAF groups. For two-group comparisons, Mann-Whitney test was used for continuous variables, and Fisher’s exact test for categorical variables. To assess if baseline levels of serum CT and other variables are predictors of POAF, logistic regression was first applied in R using the packages gtsummary (v2.0.4) and finalfit (v1.0.8) to determine if CT and other variables are independent predictors of POAF. Receiver Operating Characteristic (ROC) curve analysis was performed in R using the pROC package (v1.19.0.1) to give an area under curve (AUC) value for the logistic regression models [9]. Time-to-event analysis was performed in Prism to assess the association between baseline predictors and the onset of POAF. Cox proportional hazards regression was conducted to estimate hazard ratios (HR) for each predictor independently in the univariate analysis. Multivariate regression analyses were adjusted for covariates including sex, age, diabetes and NYHA classification. Kaplan-Meier survival curves were presented to visualise the cumulative incidence of POAF over the duration of hospitalisation. Baseline levels of serum CT were stratified by three groups according to the quartile distribution: low CT (Q1), moderate CT (Q2-3) and high CT (Q4) levels. Differences between the groups were examined using the log-rank test for trend and *P*-value < 0.05 was considered significant. Patients who remained in sinus rhythm and did not develop POAF were censored at the time of hospital discharge.

## Results

### Baseline demographic data

The study design is presented in **Figure 1**. Among 454 patients in sinus rhythm, 44 (9.7%) had missing serum CT measurements and were therefore excluded. The final study cohort consisted of 410 patients in sinus rhythm with available baseline serum CT measurements. Of these, CT levels were undetectable in 160 patients (39%). Baseline characteristics of patients with detectable and undetectable CT are presented in **Table 1**. The only significant difference between the groups was sex distribution, with a higher proportion of males in the group with detectable CT compared with those with undetectable CT (91% vs. 66%, P < 0.0001). Patients were further categorized according to whether CT levels were within the normal reference range (males: 0–11 pg/mL; females: 0–6 pg/mL pg/ml [10]) or above the reference range. The comparison between these groups is shown in **Table 2**. Patients with CT levels above the reference range were more likely to undergo CABG and had a higher prevalence of prior myocardial infarction, hypertension, and diabetes. Additionally, the proportion of patients who developed POAF was 11% lower in the group with CT levels exceeding the reference range; however, this difference did not reach statistical significance (P = 0.0817).

**Figure 1.**
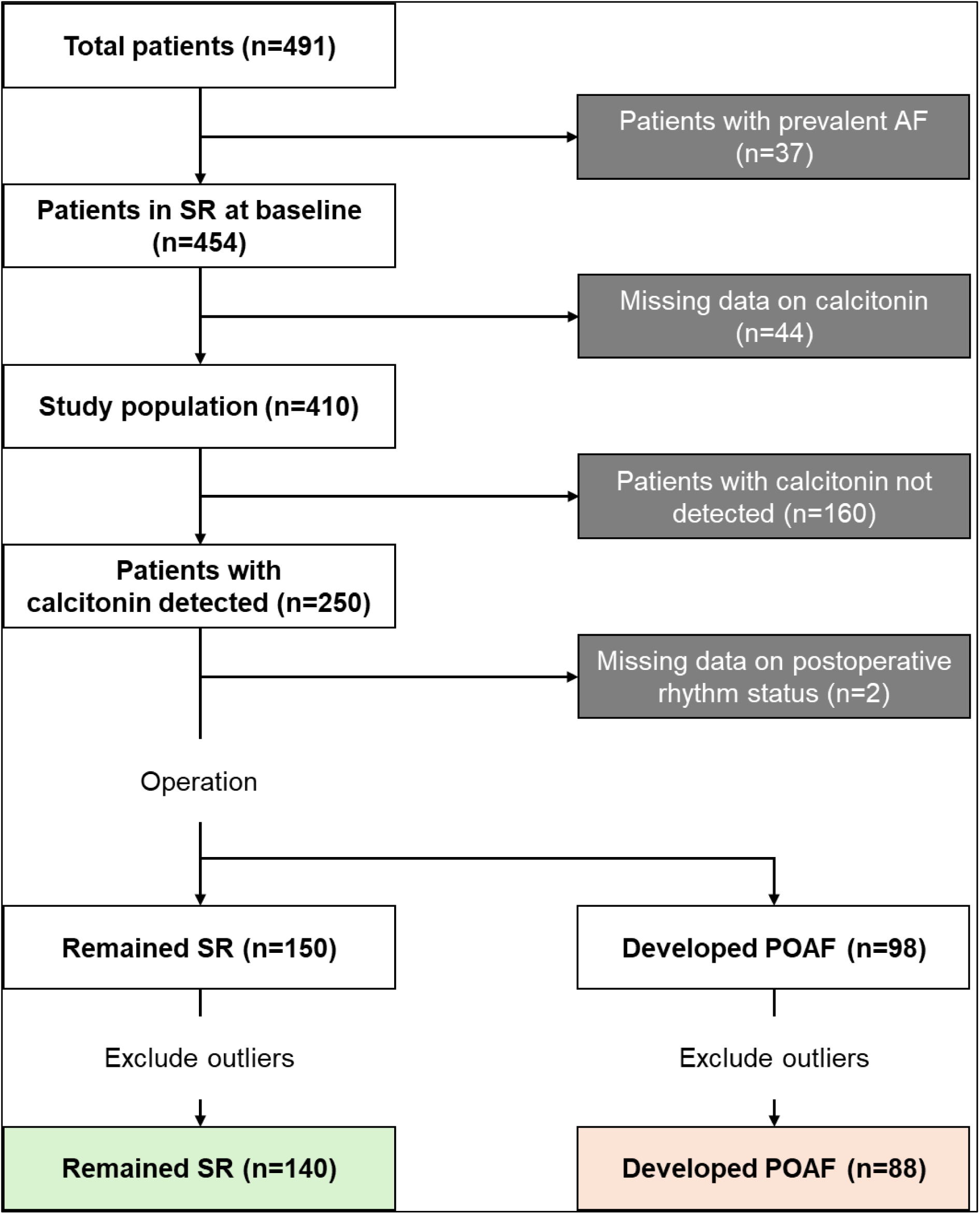
Study design. AF, atrial fibrillation; POAF, postoperative atrial fibrillation; SR, sinus rhythm.

**Table 1.** Baseline characteristics of patients with detected and non-detected calcitonin measurements. *P*-values were calculated by the Mann-Whitney test for Age and Hospitalisation, and Fisher’s exact test for other characteristics. Continuous data are median (IQR). ACEi, angiotensin-converting enzyme inhibitor; AVR, aortic valve replacement; CABG, coronary artery bypass graft; COPD, chronic obstructive pulmonary disease; CT, calcitonin; MI, myocardial infarction; ND, non-detected; NYHA, New York Heart Association; POAF, postoperative atrial fibrillation; SR, sinus rhythm.

|  | ND CT (n=160) | Detected CT (n=248) | *P*-value |
| --- | --- | --- | --- |
| **Demographic** |  |  |  |
| Male, n (%) | 105 (66) | 225 (91) | < 0.0001 |
| Age, y (IQR) | 68 (60-74) | 67 (59-75) | 0.7808 |
| **Surgical procedure** |  |  |  |
| CABG | 137 (86) | 224 (90) | 0.1556 |
| Hospitalisation, days (IQR) | 6 (5-8) | 6 (5-8) | 0.0762 |
| **Medical history, n (%)** |  |  |  |
| POAF | 63 (39) | 98 (40) | 0.9999 |
| NYHA classification |  |  |  |
| I | 73 (46) | 97 (39) | 0.2174 |
| II | 60 (38) | 104 (42) | 0.4085 |
| III | 22 (14) | 42 (17) | 0.4068 |
| IV | 5 (3) | 5 (2) | 0.5230 |
| MI | 59 (37) | 100 (40) | 0.5331 |
| Hypertension | 111 (69) | 185 (75) | 0.2577 |
| Diabetes | 32 (20) | 56 (23) | 0.6221 |
| COPD | 7 (4) | 9 (4) | 0.7956 |
| Hypo-thyroid function | 8 (5) | 8 (3) | 0.4362 |
| Hyper-thyroid function | 1 (1) | 2 (1) | 0.9999 |
| **Medication, n (%)** |  |  |  |
| Anticoagulant | 27 (17) | 41 (17) | 0.9999 |
| Antiplatelet | 134 (84) | 218 (88) | 0.2418 |
| β-blocker | 108 (68) | 175 (71) | 0.5118 |
| Statins | 138 (86) | 210 (85) | 0.7748 |
| Calcium channel blocker | 38 (24) | 70 (28) | 0.3583 |
| ACEi | 80 (50) | 122 (49) | 0.9193 |
| Diuretics | 22 (14) | 49 (20) | 0.1413 |

**Table 2.** Baseline characteristics of patients with normal and above normal calcitonin measurements. Reference ranges of calcitonin in normal physiological condition were taken from the NHS Foundation Trust (males 0-11 pg/ml; females 0-6 pg/ml). Continuous data are median (IQR). *P*-values were calculated by the Mann-Whitney test for Age and Hospitalisation, and Fisher’s exact test for other characteristics. ACEi, angiotensin-converting enzyme inhibitor; AVR, aortic valve replacement; CABG, coronary artery bypass graft; COPD, chronic obstructive pulmonary disease; CT, calcitonin; MI, myocardial infarction; NYHA, New York Heart Association; POAF, postoperative atrial fibrillation; SR, sinus rhythm.

|  | Normal (n=338) | Above normal (n=70) | *P*-value |
| --- | --- | --- | --- |
| **Demographic** |  |  |  |
| Male, n (%) | 269 (80) | 61 (87) | 0.1813 |
| Age, y (IQR) | 68 (60-75) | 65 (59-72) | 0.1281 |
| **Surgical procedure** |  |  |  |
| CABG | 293 (87) | 68 (97) | 0.0120 |
| Hospitalisation, days (IQR) | 6 (5-8) | 6 (5-9) | 0.4664 |
| **Medical history, n (%)** |  |  |  |
| POAF | 140 (41) | 21 (30) | 0.0817 |
| NYHA classification |  |  |  |
| I | 142 (42) | 28 (40) | 0.7912 |
| II | 135 (40) | 29 (41) | 0.8936 |
| III | 53 (16) | 11 (16) | 0.9999 |
| IV | 8 (2) | 2 (3) | 0.6835 |
| MI | 122 (36) | 37 (53) | 0.0105 |
| Hypertension | 235 (70) | 61 (87) | 0.0020 |
| Diabetes | 62 (18) | 26 (37) | 0.0012 |
| COPD | 13 (4) | 3 (4) | 0.7440 |
| Hypo-thyroid function | 13 (4) | 3 (4) | 0.7440 |
| Hyper-thyroid function | 1 (0) | 2 (3) | 0.0775 |
| **Medication, n (%)** |  |  |  |
| Anticoagulant | 51 (15) | 17 (24) | 0.0768 |
| Antiplatelet | 287 (85) | 65 (93) | 0.0872 |
| β-blocker | 229 (68) | 54 (77) | 0.1539 |
| Statins | 286 (85) | 62 (89) | 0.4624 |
| Calcium channel blocker | 83 (25) | 25 (36) | 0.0731 |
| ACEi | 165 (49) | 37 (53) | 0.5999 |
| Diuretics | 58 (17) | 13 (19) | 0.7324 |

Postoperative rhythm status was assessed for each patient; however, data were missing for 2 patients. Therefore, the analysis included the remaining 248 patients with detectable baseline serum CT levels, comprising 150 patients who remained in sinus rhythm and 88 patients who developed POAF. Given the variability of CT levels within groups, the Robust Regression and Outlier Removal (ROUT) test with Q = 0.1% was applied to identify and exclude outliers. Following this procedure, 140 patients were included in the sinus rhythm group and 88 patients in the POAF group. Baseline characteristics based on postoperative rhythm status are summarised in **Table 3**. Compared to patients who remained in sinus rhythm, POAF patients were older (*P* < 0.0001), longer days of hospitalisation (*P* < 0.0001) and lower proportion of patients classified as NYHA Class I (45% vs. 30%, *P* = 0.0255). No differences were observed between groups regarding the medications.

**Table 3.** Baseline characteristics of patients with calcitonin measurements. Continuous data are median (IQR). *P*-values were calculated by the Mann-Whitney test for Age and Hospitalisation, and Fisher’s exact test for other characteristics. ACEi, angiotensin-converting enzyme inhibitor; CABG, coronary artery bypass graft; COPD, chronic obstructive pulmonary disease; CT, calcitonin; MI, myocardial infarction; NYHA, New York Heart Association; POAF, postoperative atrial fibrillation; SR, sinus rhythm.

| Characteristic | SR (n=140) | POAF (n=88) | *P*-value |
| --- | --- | --- | --- |
| **Demographic** |  |  |  |
| Male, n (%) | 124 (89) | 83 (94) | 0.1650 |
| Age, y (IQR) | 64 (57-72) | 71 (65-76) | < 0.0001 |
| **Surgical procedure** |  |  |  |
| CABG | 128 (91) | 76 (86) | 0.2693 |
| Hospitalisation, days (IQR) | 6 (5-7) | 7 (6-10) | < 0.0001 |
| **Medical history, n (%)** |  |  |  |
| NYHA classification |  |  |  |
| I | 63 (45) | 26 (30) | 0.0255 |
| II | 53 (38) | 43 (49) | 0.1295 |
| III | 20 (14) | 18 (20) | 0.2737 |
| IV | 4 (3) | 1 (1) | 0.6512 |
| MI | 54 (39) | 35 (40) | 0.8896 |
| Hypertension | 100 (71) | 69 (78) | 0.2784 |
| Diabetes | 36 (26) | 13 (15) | 0.0678 |
| COPD | 4 (3) | 5 (6) | 0.3122 |
| Hypo-thyroid function | 5 (4) | 3 (3) | 0.9999 |
| Hyper-thyroid function | 1 (1) | 0 (0) | 0.9999 |
| **Medication, n (%)** |  |  |  |
| Anticoagulant | 27 (19) | 12 (14) | 0.2858 |
| Antiplatelet | 124 (89) | 74 (84) | 0.4211 |
| β-blocker | 102 (73) | 58 (66) | 0.2989 |
| Statins | 120 (86) | 72 (82) | 0.4592 |
| Calcium channel blocker | 38 (27) | 24 (27) | 0.9999 |
| ACEi | 72 (51) | 41 (47) | 0.4988 |
| Diuretics | 28 (30) | 20 (23) | 0.6212 |

### Baseline serum CT associates with the risk of POAF

Logistic regression model indicated that each 5 pg/mL increase in baseline serum CT was associated with a lower risk of POAF [OR 0.69 (95% CI 0.52–0.88), P = 0.004] (**Table 4**). Increasing age and NYHA class II (compared to class I) remained as independent predictors of higher risk of POAF in univariate analysis. After adjusting for covariates including sex, age, NYHA class, and diabetes, each 5 pg/mL increase in baseline serum CT associated with a lower risk of POAF [OR 0.68 (95% CI 0.51–0.89), P = 0.009], and increasing age remained an independent predictor of POAF [OR 2.03 (95% CI 1.47–2.89) per 10-year increase, P < 0.001] (**Table 5** and **Figure 2a**). Receiver operating characteristic (ROC) curves for the univariate and multivariable logistic regression models showed AUC values of 0.5896 and 0.7392, respectively (**Figure 2b**). Blood biomarkers associated with POAF include C-reactive protein (CRP) and plasma B-type natriuretic peptide (BNP) [11]; however, their baseline levels were not associated with the risk of developing POAF in this study cohort. In contrast to these traditional biomarkers, baseline serum CT independently predicted the risk of POAF.

**Figure 2.**
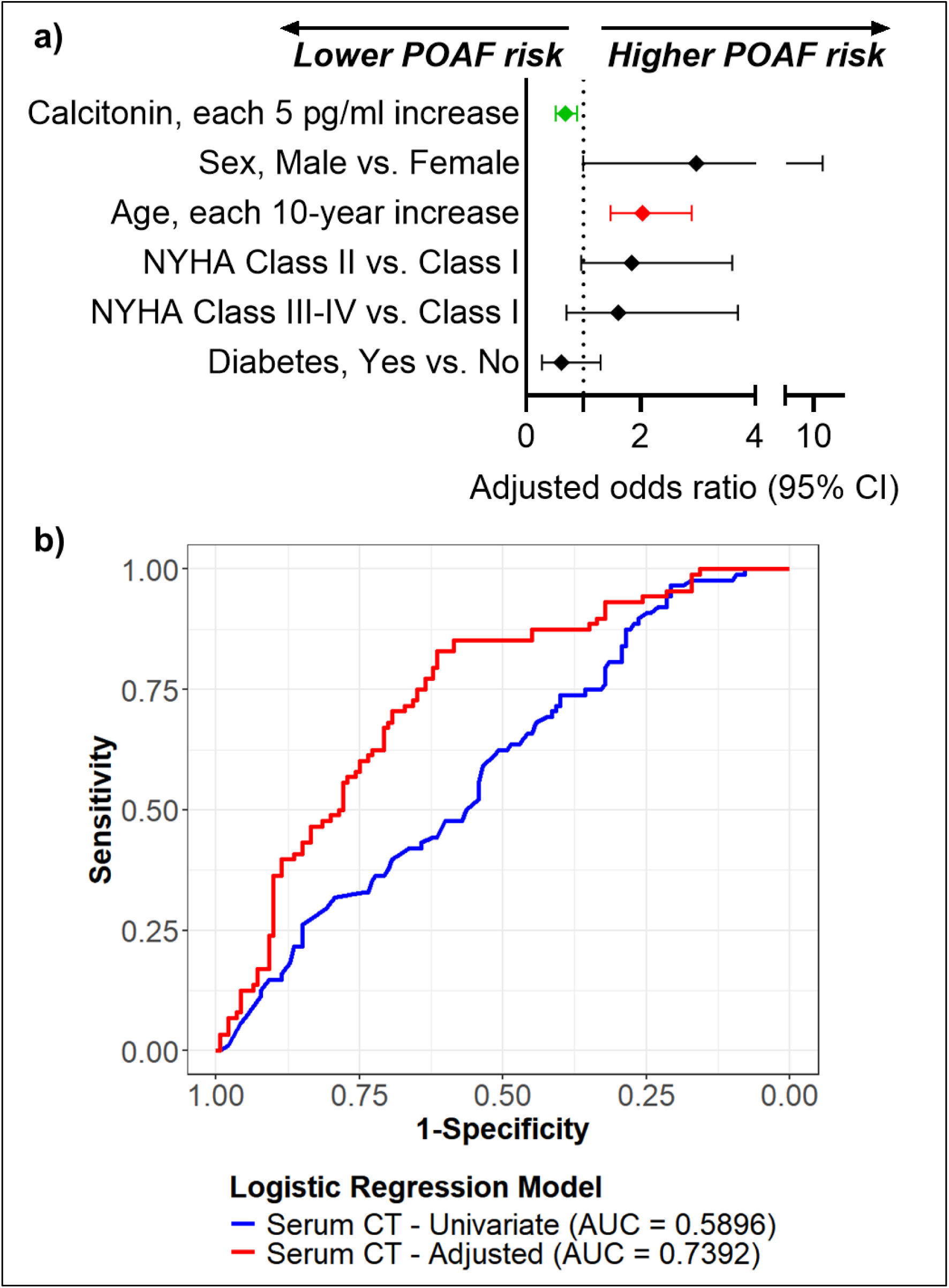
High concentrations of CT at baseline predict lower risk of POAF after cardiac surgery. (**a**) Forest plot showing odds ratios and 95% confidence intervals for predictors of POAF in the multivariate logistic regression model. (**b**) ROC curves showing the performance of univariate and multivariate models in predicting POAF risk. AUC, area under curve; CI, confidence interval; HR, hazard ratio; NYHA, New York Heart Association; OR, odds ratio; POAF, postoperative atrial fibrillation; ROC, receiver operating characteristic curve; SR, sinus rhythm.

**Table 4.** Univariate logistic regression analysis of baseline serum calcitonin for predicting postoperative atrial fibrillation risk. Characteristics with * indicate that data were not available in all patients. ACEi, angiotensin-converting enzyme inhibitor; CABG, coronary artery bypass graft; CI, confidence interval; HR, hazard ratio; MI, myocardial infarction; NYHA, New York Heart Association; POAF, postoperative atrial fibrillation; SR, sinus rhythm.

| Characteristic |  |  | SR | POAF | Univariate |  |  |
| --- | --- | --- | --- | --- | --- | --- | --- |
|  |  |  |  |  | OR | 95% CI | P value |
| Serum calcitonin (pg/ml) | For each 5 pg/ml increase | Median (IQR) | 5.0 (2.1-11.7) | 4.3 (1.9-7.9) | 0.69 | 0.52-0.88 | 0.004 |
| Sex |  | Female | 16 (11.4) | 5 (5.7) |  |  |  |
|  | Male vs. Female | Male | 124 (88.6) | 83 (94.3) | 2.14 | 0.80-6.75 | 0.152 |
| Age | for each 10-year increase | Median (IQR) | 64 (57-72) | 71 (65-76) | 1.99 | 1.47-2.76 | < 0.001 |
| CABG |  | No | 12 (8.6) | 12 (13.6) |  |  |  |
|  | Yes vs. No | Yes | 128 (91.4) | 76 (86.4) | 0.59 | 0.25-1.40 | 0.229 |
| Diabetes |  | No | 104 (74.3) | 75 (85.2) |  |  |  |
|  | Yes vs. No | Yes | 36 (25.7) | 13 (14.8) | 0.50 | 0.24-0.99 | 0.053 |
| NYHA classification |  | I | 63 (45.0) | 26 (29.5) |  |  |  |
|  | II vs. I | II | 53 (37.9) | 43 (48.9) | 1.97 | 1.08-3.64 | 0.030 |
|  | III-IV vs. I | III-IV | 24 (17.1) | 19 (21.6) | 1.92 | 0.90-4.10 | 0.091 |
| MI |  | No | 86 (61.4) | 53 (60.2) |  |  |  |
|  | Yes vs. No | Yes | 54 (38.6) | 35 (39.8) | 1.05 | 0.61-1.81 | 0.856 |
| Hypertension |  | No | 40 (28.6) | 19 (21.6) |  |  |  |
|  | Yes vs. No | Yes | 100 (71.4) | 69 (78.4) | 1.45 | 0.78-2.76 | 0.243 |
| Anticoagulant |  | No | 113 (80.7) | 76 (86.4) |  |  |  |
|  | Yes vs. No | Yes | 27 (19.3) | 12 (13.6) | 0.66 | 0.31-1.36 | 0.272 |
| Antiplatelet |  | No | 16 (11.4) | 14 (15.9) |  |  |  |
|  | Yes vs. No | Yes | 124 (88.6) | 74 (84.1) | 0.68 | 0.31-1.49 | 0.332 |
| β-blocker |  | No | 38 (27.1) | 30 (34.1) |  |  |  |
|  | Yes vs. No | Yes | 102 (72.9) | 58 (65.9) | 0.72 | 0.40-1.29 | 0.265 |
| Statins |  | No | 20 (14.3) | 16 (18.2) |  |  |  |
|  | Yes vs. No | Yes | 120 (85.7) | 72 (81.8) |  |  |  |
| Calcium channel blocker |  | No | 102 (72.9) | 64 (72.7) | 0.75 | 0.37-1.56 | 0.433 |
|  | Yes vs. No | Yes | 38 (27.1) | 24 (27.3) | 1.01 | 0.55-1.82 | 0.983 |
| ACEi |  | No | 68 (48.6) | 47 (53.4) |  |  |  |
|  | Yes vs. No | Yes | 72 (51.4) | 41 (46.6) | 0.82 | 0.48-1.40 | 0.477 |
| Diuretics |  | No | 112 (80.0) | 68 (77.3) |  |  |  |
|  | Yes vs. No | Yes | 28 (20.0) | 20 (22.7) | 1.18 | 0.61-2.24 | 0.623 |
| *White blood count (cells/μl) | for each 1000 cells/μl increase | Mean (SD) | 7682.7 (3467.8) | 7120.1 (1772.0) | 0.92 | 0.80-1.03 | 0.187 |
| *Hemoglobin |  | Mean (SD) | 14.0 (1.4) | 13.9 (1.1) | 0.92 | 0.75-1.14 | 0.463 |
| *CRP |  | Mean (SD) | 3.0 (4.9) | 3.7 (7.4) | 1.02 | 0.96-1.08 | 0.453 |
| *Plasma BNP (pg/ml) | for each 10 pg/ml increase | Mean (SD) | 85.5 (154.8) | 118.4 (168.4) | 1.01 | 0.99-1.03 | 0.176 |
| *Creatinine (μmol/l) | for each 10 μmol/l increase | Mean (SD) | 85.3 (28.1) | 87.5 (22.1) | 1.03 | 0.93-1.15 | 0.560 |
| *LVEF |  | Good (>50%) | 120 (86.3) | 71 (82.6) |  |  |  |
| | | Reduced (≤50%) | 19 (13.7) | 15 (17.4) | 1.33 | 0.63-2.79 | 0.444 |
| *E/A |  | Mean (SD) | 1.0 (1.0) | 1.0 (0.5) | 1.01 | 0.62-1.53 | 0.979 |
| *LA length |  | Mean (SD) | 5.3 (0.8) | 5.5 (0.8) | 1.29 | 0.67-2.61 | 0.452 |
| *LA diameter |  | Mean (SD) | 3.9 (0.6) | 3.9 (0.7) | 0.98 | 0.41-2.30 | 0.966 |
| *LVEDD |  | Mean (SD) | 4.8 (0.6) | 5.0 (0.8) | 1.40 | 0.93-2.11 | 0.108 |
| *LVEDV |  | Mean (SD) | 3.1 (0.8) | 3.2 (1.0) | 1.09 | 0.74-1.63 | 0.653 |
| *LVESD | for each 10 units increase | Mean (SD) | 108.6 (39.8) | 121.0 (48.9) | 1.07 | 0.98-1.16 | 0.132 |
| *LVESV | for each 10 units increase | Mean (SD) | 44.5 (25.6) | 52.7 (39.8) | 1.08 | 0.96-1.22 | 0.189 |

**Table 5.** Multivariate logistic regression analysis of baseline serum calcitonin for predicting postoperative atrial fibrillation risk. CI, confidence interval; HR, hazard ratio; NYHA, New York Heart Association; POAF, postoperative atrial fibrillation; SR, sinus rhythm.

| Characteristic |  |  | SR | POAF | Multivariate |  |  |
| --- | --- | --- | --- | --- | --- | --- | --- |
|  |  |  |  |  | OR | 95% CI | P value |
| Serum calcitonin (pg/ml) | for each 5 pg/ml increase | Median (IQR) | 5.0 (2.1-11.7) | 4.3 (1.9-7.9) | 0.68 | 0.51-0.89 | 0.009 |
| Sex |  | Female | 16 (11.4) | 5 (5.7) |  |  |  |
|  | Male vs. Female | Male | 124 (88.6) | 83 (94.3) | 2.97 | 0.99-10.30 | 0.064 |
| Age | for each 10-year increase | Median (IQR) | 64 (57-72) | 71 (65-76) | 2.03 | 1.47-2.89 | < 0.001 |
| Diabetes |  | No | 104 (74.3) | 75 (85.2) |  |  |  |
|  | Yes vs. No | Yes | 36 (25.7) | 13 (14.8) | 0.61 | 0.27-1.30 | 0.206 |
| NYHA classification |  | I | 63 (45.0) | 26 (29.5) |  |  |  |
|  | II vs. I | II | 53 (37.9) | 43 (48.9) | 1.84 | 0.96-3.60 | 0.070 |
|  | III-IV vs. I | III-IV | 24 (17.1) | 19 (21.6) | 1.61 | 0.70-3.70 | 0.257 |

### Higher baseline serum CT associates with delayed POAF onset

In time-to-event analysis using the Cox proportional hazards model (**Table 6** and **Figure 3a**), each 1 pg/mL increase in baseline serum CT was associated with a 6% lower instantaneous hazard of developing POAF [HR 0.944 (95% CI 0.902–0.983), P = 0.0045]. After adjustment for covariates including sex, age, NYHA class, and diabetes, this association remained significant and of similar magnitude [adjusted HR 0.941 (95% CI 0.898–0.980), P = 0.0026] (**Table 7**). Kaplan–Meier survival analysis was performed to evaluate the time to onset of POAF across tertiles of baseline serum CT. Patients were categorised into three groups: low CT (Q1), medium CT (Q2–Q3), and high CT (Q4). The curves indicate that patients with higher baseline serum CT levels experienced a delayed or lower incidence of POAF onset (**Figure 3b**). A log-rank test for trend demonstrated a statistically significant difference in time-to-event distributions across the three groups (χ² = 5.290, P = 0.0214). These findings support the potential relevance of baseline serum CT as a cardioprotective marker for atrial remodelling. In addition to POAF, other postoperative and long-term clinical outcomes were evaluated (**Table 8**) and showed no association with baseline serum levels of CT, further supporting the specificity of CT for atrial rhythm outcomes rather than general postoperative complications.

**Figure 3.**
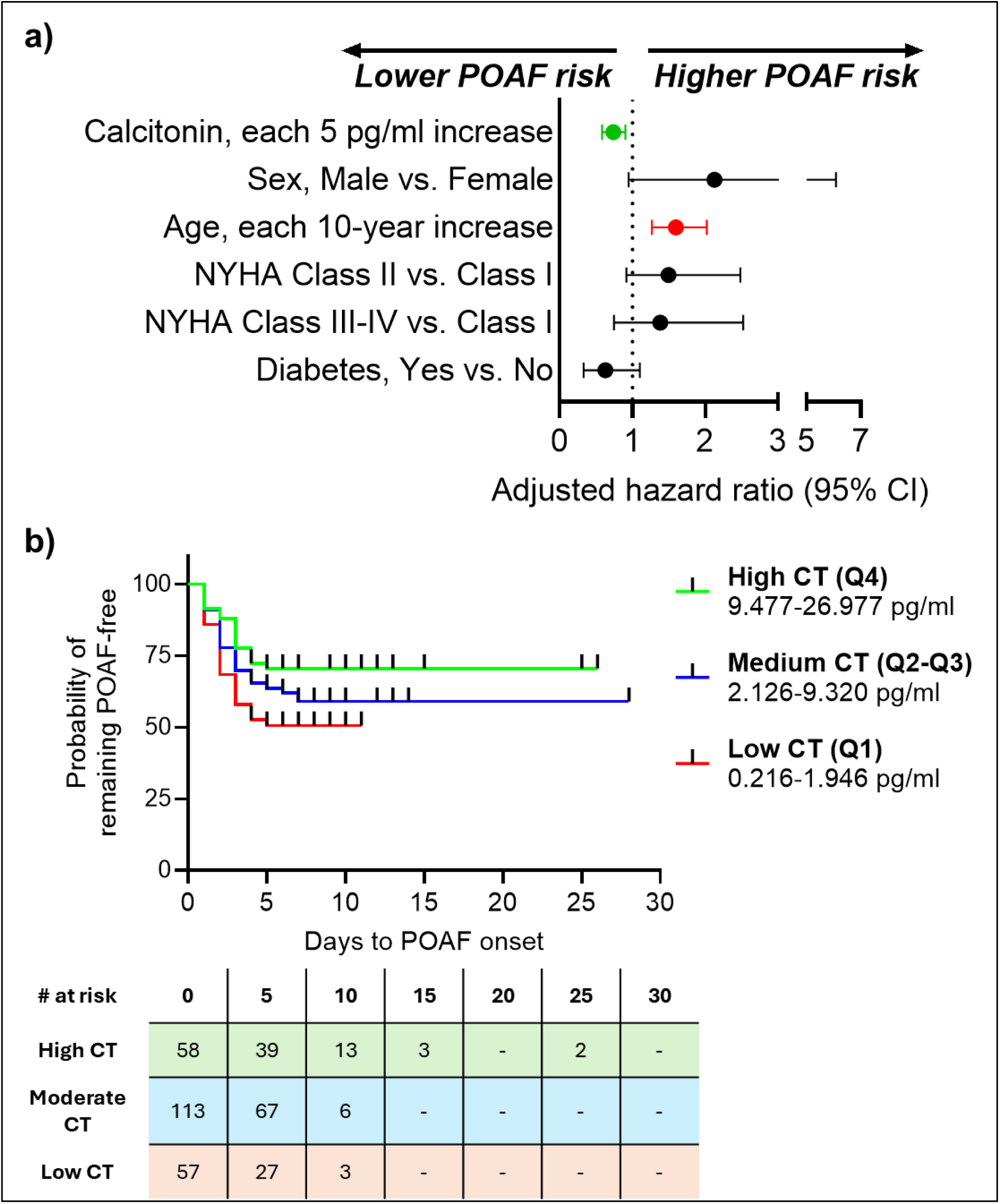
Patients with higher concentrations of CT at baseline have delayed onset for POAF. (**a**) (**a**) Forest plot showing hazard ratios and 95% confidence intervals for predictors of POAF in the multivariate Cox proportional hazards regression model. (**b**) Kaplan-Meier curves showing the probabilities of remaining POAF-free after cardiac surgery in three groups with different CT levels: low CT (Q1, lowest 25%), medium CT (Q2-Q3, 25%-75%) and high CT (Q4, highest 25%). CI, confidence interval; CT, calcitonin; HR, hazard ratio; POAF, postoperative atrial fibrillation.

**Table 6.**
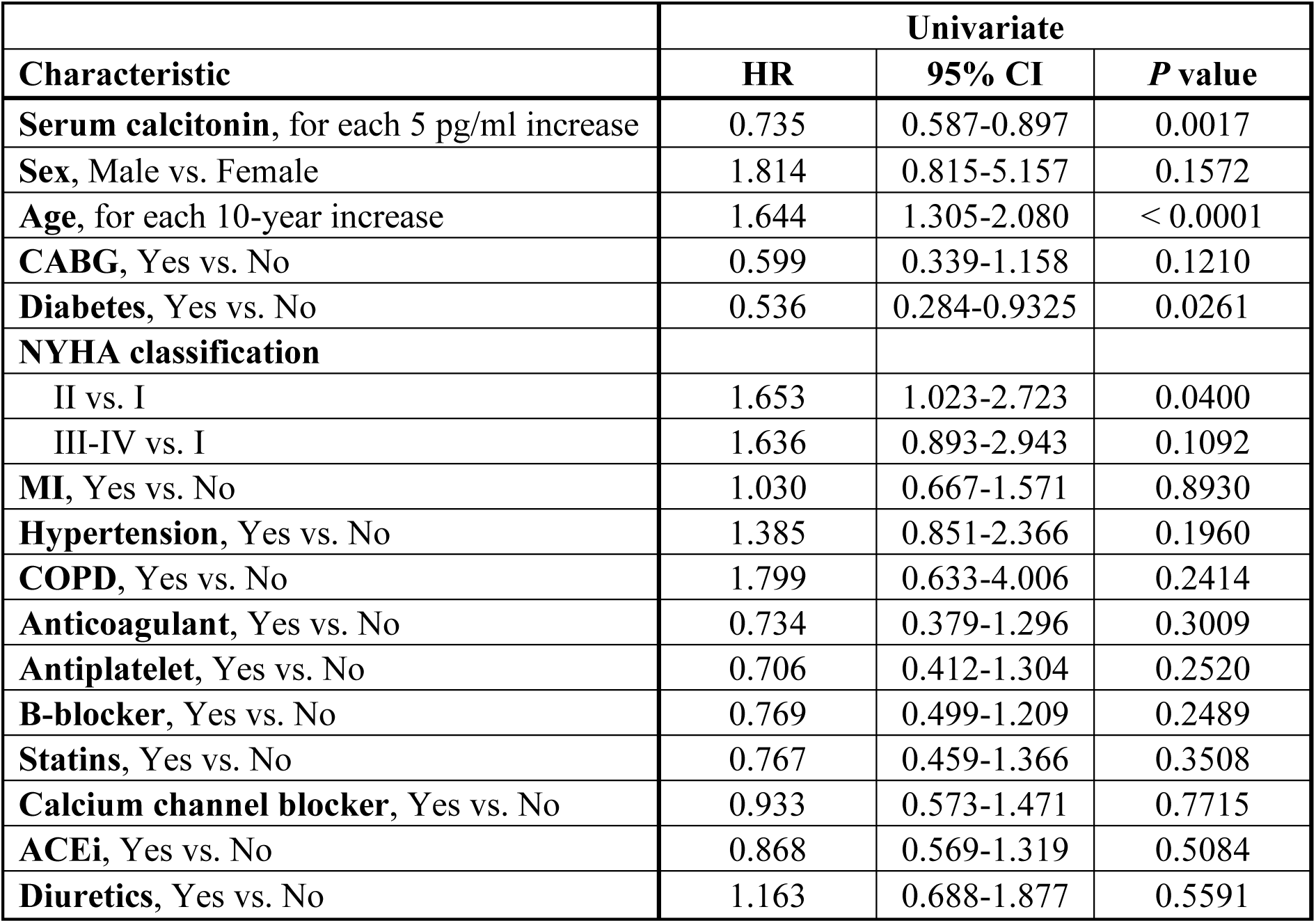
Cox proportional hazard regression univariate analysis of baseline serum calcitonin for predicting postoperative atrial fibrillation risk. ACEi, angiotensin-converting enzyme inhibitor; CABG, coronary artery bypass graft; CI, confidence interval; COPD, chronic obstructive pulmonary disease; HR, hazard ratio; MI, myocardial infarction; NYHA, New York Heart Association; POAF, postoperative atrial fibrillation; SR, sinus rhythm.

| Characteristic | Univariate |  |  |
| --- | --- | --- | --- |
|  | HR | 95% CI | P value |
| **Serum calcitonin**, for each 5 pg/ml increase | 0.735 | 0.587-0.897 | 0.0017 |
| **Sex**, Male vs. Female | 1.814 | 0.815-5.157 | 0.1572 |
| **Age**, for each 10-year increase | 1.644 | 1.305-2.080 | < 0.0001 |
| **CABG**, Yes vs. No | 0.599 | 0.339-1.158 | 0.1210 |
| **Diabetes**, Yes vs. No | 0.536 | 0.284-0.9325 | 0.0261 |
| **NYHA classification** |  |  |  |
| II vs. I | 1.653 | 1.023-2.723 | 0.0400 |
| III-IV vs. I | 1.636 | 0.893-2.943 | 0.1092 |
| **MI**, Yes vs. No | 1.030 | 0.667-1.571 | 0.8930 |
| **Hypertension**, Yes vs. No | 1.385 | 0.851-2.366 | 0.1960 |
| **COPD**, Yes vs. No | 1.799 | 0.633-4.006 | 0.2414 |
| **Anticoagulant**, Yes vs. No | 0.734 | 0.379-1.296 | 0.3009 |
| **Antiplatelet**, Yes vs. No | 0.706 | 0.412-1.304 | 0.2520 |
| **B-blocker**, Yes vs. No | 0.769 | 0.499-1.209 | 0.2489 |
| **Statins**, Yes vs. No | 0.767 | 0.459-1.366 | 0.3508 |
| **Calcium channel blocker**, Yes vs. No | 0.933 | 0.573-1.471 | 0.7715 |
| **ACEi**, Yes vs. No | 0.868 | 0.569-1.319 | 0.5084 |
| **Diuretics**, Yes vs. No | 1.163 | 0.688-1.877 | 0.5591 |

**Table 7.** Cox proportional hazard regression multivariate analysis of baseline serum calcitonin for predicting postoperative atrial fibrillation risk. CI, confidence interval; HR, hazard ratio; NYHA, New York Heart Association; POAF, postoperative atrial fibrillation; SR, sinus rhythm.

| **Characteristic** | **Multivariate** |  |  |
| --- | --- | --- | --- |
|  | **HR** | **95% CI** | **P value** |
| **Serum calcitonin**, for each 5 pg/ml increase | 0.737 | 0.585-0.904 | 0.0026 |
| **Sex**, Male vs. Female | 2.128 | 0.948-6.084 | 0.0695 |
| **Age**, for each 10-year increase | 1.598 | 1.268-2.023 | < 0.0001 |
| **Diabetes**, Yes vs. No | 0.630 | 0.332-1.103 | 0.0936 |
| **NYHA classification** |  |  |  |
| II vs. I | 1.497 | 0.919-2.483 | 0.1055 |
| III-IV vs. I | 1.384 | 0.747-2.522 | 0.2962 |

**Table 8.**
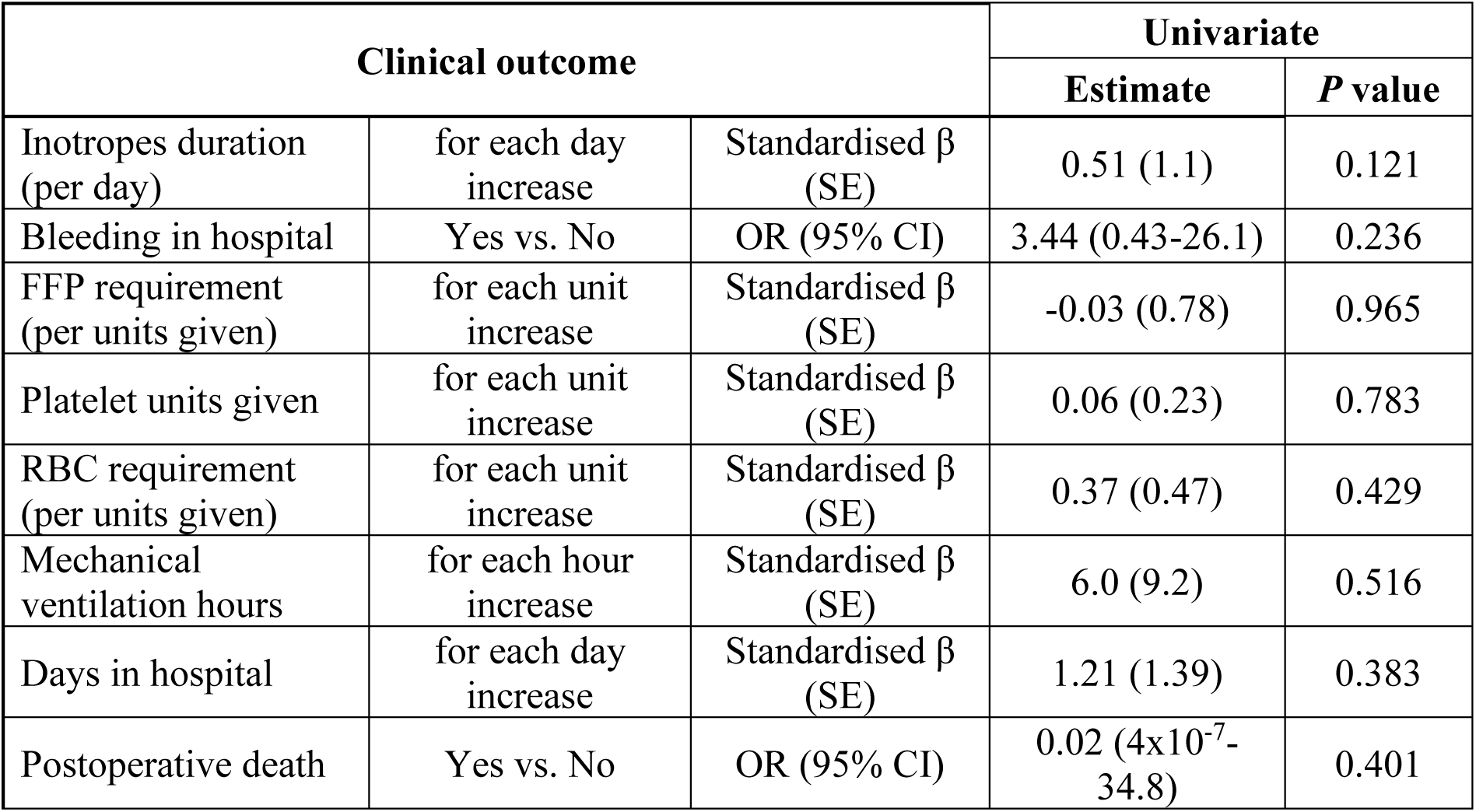
Association of baseline serum calcitonin with postoperative clinical outcomes. Linear regression was used for continuous outcomes (log-transformed). Logistic regression was used for other categorical outcomes. AF, atrial fibrillation; CI, confidence interval; FFP, fresh frozen plasma; NYHA, New York Heart Association; OR, odds ratio; POAF, postoperative atrial fibrillation; RBC, red blood cell; SE, standard error; SR, sinus rhythm.

| Clinical outcome |  |  | Univariate |  |
| --- | --- | --- | --- | --- |
|  |  |  | Estimate | P value |
| Inotropes duration (per day) | for each day increase | Standardised $\beta$ (SE) | 0.51 (1.1) | 0.121 |
| Bleeding in hospital | Yes vs. No | OR (95% CI) | 3.44 (0.43-26.1) | 0.236 |
| FFP requirement (per units given) | for each unit increase | Standardised $\beta$ (SE) | -0.03 (0.78) | 0.965 |
| Platelet units given | for each unit increase | Standardised $\beta$ (SE) | 0.06 (0.23) | 0.783 |
| RBC requirement (per units given) | for each unit increase | Standardised $\beta$ (SE) | 0.37 (0.47) | 0.429 |
| Mechanical ventilation hours | for each hour increase | Standardised $\beta$ (SE) | 6.0 (9.2) | 0.516 |
| Days in hospital | for each day increase | Standardised $\beta$ (SE) | 1.21 (1.39) | 0.383 |
| Postoperative death | Yes vs. No | OR (95% CI) | 0.02 ($4 \times 10^{-7}$-34.8) | 0.401 |

## Discussion

Our study demonstrates that preoperative circulating CT independently predicts a lower incidence and delayed onset of POAF following cardiac surgery. Despite the immense clinical burden of POAF, which affects up to 55% of cardiac surgery patients and drives significant long-term morbidity [1, 2, 4], current clinical risk prediction models have only modest discriminatory ability [5, 6]. In our cohort, 39.5% of patients developed POAF, which is consistent with previously reported incidences in mixed cardiac surgery populations [1, 2]. Our findings directly address this gap. We extend the current knowledge of POAF pathophysiology by linking the known experimental cardioprotective properties of CT to acute clinical outcomes, identifying circulating CT as a novel biomarker with potential utility for perioperative risk stratification.

To our knowledge, this study is the first to evaluate circulating CT as a predictor of POAF. While previous research has focused on CT in endocrine and systemic diseases, its role in acute cardiovascular arrhythmia risk has not been explored [22]. Unlike traditional biomarkers such as C-reactive protein (CRP) and B-type natriuretic peptide (BNP), which did not predict POAF in our cohort, baseline CT independently predicted risk and may reflect underlying pathophysiological processes related to atrial remodelling. Other circulating markers, including N-terminal pro-brain natriuretic peptide (NT-proBNP), matrix metalloproteinase-9 (MMP9), and interleukin-6 (IL-6), have been identified as modest predictors of POAF, but circulating CT offers the added advantage of being easily accessible, minimally invasive, and well-suited for perioperative risk stratification. These findings extend the relevance of CT to acute postoperative outcomes and highlight its potential translational utility in clinical practice.

POAF is widely recognized as a multifactorial condition, driven heavily by systemic inflammatory activation secondary to surgical trauma, cardiopulmonary bypass, and ischemia-reperfusion injury [3]. While elevated generic inflammatory markers have historically been associated with POAF, traditional stress and acute-phase biomarkers such as C-reactive protein (CRP) and B-type natriuretic peptide (BNP) failed to predict POAF in our cohort [11]. In contrast, baseline CT independently predicted POAF risk and was strictly associated with atrial rhythm outcomes, rather than generalized postoperative complications such as excessive bleeding, prolonged mechanical ventilation, or extended hospital length of stay (**Table 8**). This biological specificity underscores that CT is not simply a passive proxy for generalized surgical vulnerability. Our results position CT as a potentially superior biomarker, capturing distinct atrial electrophysiological processes that are not reflected by generic systemic inflammation or natriuretic signalling alone.

One notable finding in our cohort was the distinct clinical phenotype of patients presenting with above-normal baseline CT levels (**Table 2**). Counterintuitively, these individuals carried a considerably higher burden of severe cardiovascular and metabolic comorbidities, including a history of myocardial infarction, hypertension, and a greater need for CABG procedures. Furthermore, we found that patients with elevated CT had a significantly higher prevalence of diabetes (37% vs. 18%). The relationship between diabetes and POAF remains controversial, with studies reporting increased, decreased, or neutral associations [1, 3, 19–21]. Despite this more severe baseline cardiovascular risk profile, these patients demonstrated a trend toward lower susceptibility to POAF. While speculative, it may be that elevated circulating CT in this high-risk population may reflect a cardioprotective response to a longstanding burden of cardiovascular and metabolic disease, potentially acting as an endogenous buffer that pre-conditions the atria and attenuates the pro-arrhythmic impact of acute surgical stress. The observed association with diabetes is intriguing but remains hypothesis-generating, and the potential link between metabolic state, therapies, and CT upregulation requires validation in future studies.

Acute triggers play a major role in the pathogenesis of POAF with the highest vulnerability occurring early after surgery, peaking around postoperative day 2 [3]. Experimental evidence demonstrates that cardiac CT exerts potent anti-fibrotic effects by limiting atrial structural remodelling [8]. The temporal shift in arrhythmia onset observed in our cohort raises the possibility that this pathway may warrant further investigation in postoperative setting.ll [23–25]. Patients with higher baseline CT may help refine perioperative risk stratification strategies.

Consistent with the existing literature, advancing age emerged as the strongest clinical predictor of POAF, reflecting age-related structural and electrophysiological degeneration [6, 15]. Crucially, several studies have reported that serum CT levels decline with age [16–18], and CT expression in human atrial cardiomyocytes negatively correlates with age [8]. Therefore, an age-associated downregulation of CT and the subsequent loss of this cardioprotective buffer may be associated with increased susceptibility to POAF observed in older patients. Additionally, while a higher proportion of male patients had detectable CT levels, consistent with known sex-related hormonal differences [12–14], our analyses restricted to patients with measurable CT demonstrated a clear inverse association between absolute baseline CT levels and POAF risk. These findings raise the possibility that sex-specific baseline CT measurement could inform personalized perioperative risk assessment, highlighting that quantitative variation within the detectable range, rather than mere detectability, dictates biological relevance and risk stratification.

Our study included 248 patients and was conducted in a single centre; therefore, the findings should be validated in larger multi-centre cohorts. As an observational study, our results demonstrate associations rather than causal relationships. Integration of additional biomarkers, such as NT-proBNP, carboxyl terminal propeptide of collagen I, MMP9 and IL-6 [11] may further improve POAF risk prediction in high-risk patients. In addition, patients with calcitonin concentrations below the assay detection threshold were excluded from the analyses, which may have introduced a degree of selection bias; however, this approach was necessary to ensure analytical reliability and robustness of the biomarker-based modelling.

In summary, circulating CT emerges as a novel biomarker associated with POAF, with potential relevance for perioperative risk stratification in patients undergoing cardiac surgery. Importantly, circulating CT offers substantial translational potential, as it is easily accessible, minimally invasive, and ideally suited for routine clinical practice. By capturing a specific pathophysiological axis that traditional biomarkers miss, quantitative CT measurement can significantly enhance current perioperative risk assessment frameworks. Although preliminary, these findings support emerging mechanistic evidence linking CT signalling to atrial electrophysiology and raise the possibility that targeted modulation of this pathway could offer a novel therapeutic avenue for POAF prevention.

## Acknowledgements

We would like to thank all participants who provided blood samples, clinical team and investigators, especially Annette Burgess, contributing to the data generation.

## Data availability

Anonymised raw data may be provided upon reasonable request to corresponding author.

## Supplementary figure

**Figure S1.**
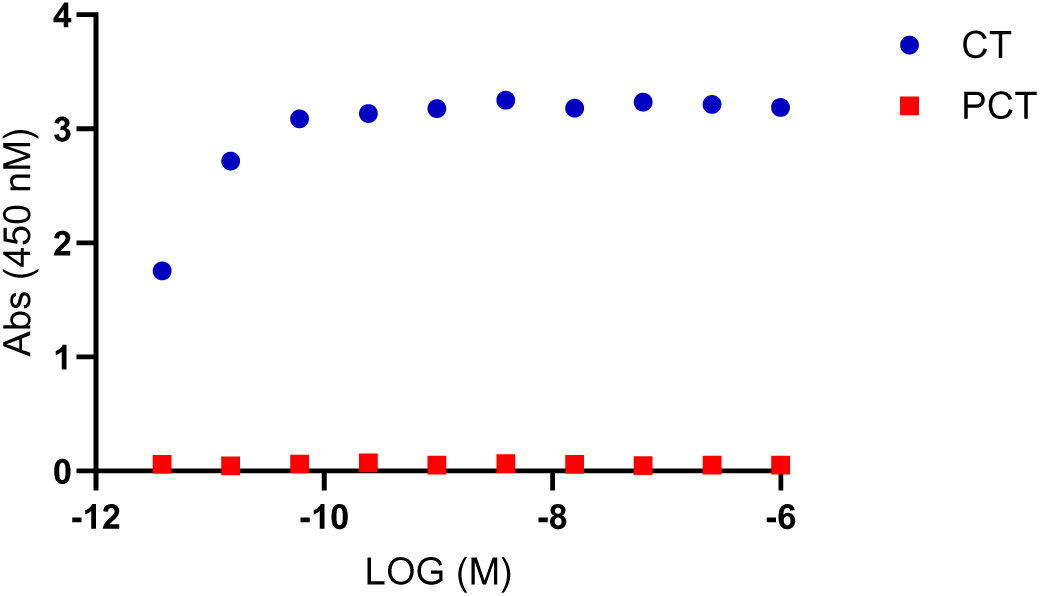
Specificity of the BioVendor serum calcitonin ELISA kit. Recombinant calcitonin (CT; blue circles) and procalcitonin (PCT; red squares) were tested across a range of concentrations using serum CT ELISA kit (BioVendor). CT produced a robust, concentration-dependent signal, whereas PCT generated no detectable signal above background across the same concentration range. These data confirm that the assay specifically detects CT and does not cross-react with PCT.

